# Systemic inflammation impairs human myelopoiesis and interferon I responses

**DOI:** 10.1101/2022.10.06.22280312

**Authors:** Farid Keramati, Guus P. Leijte, Niklas Bruse, Inge Grondman, Ehsan Habibi, Cristian Ruiz-Moreno, Wout Megchelenbrink, Annemieke M. Peters van Ton, Hidde Heesakkers, Manita Bremmers, Erinke van Grinsven, Kiki Tesselaar, Selma van Staveren, Walter van der Velden, Frank Preijers, Jelle Gerretsen, Mihai G. Netea, Hendrik G. Stunnenberg, Peter Pickkers, Matthijs Kox

## Abstract

Systemic inflammation (SI) plays a detrimental role in various conditions with high mortality rates^1–4^. SI manifests an acute hyperinflammation followed by long-lasting immunosuppression, increasing patients’ risks for secondary infections and impaired clinical outcomes^5–7^. Due to the extensive heterogeneity in SI etiology, the mechanisms governing these states are incompletely understood. Here, we characterized acute and late effects of lipopolysaccharide (LPS)-induced SI (LPS-SI^8^) on blood monocytes and bone marrow (BM) cells of healthy volunteers. Like clinical SI, LPS administration elicited a profound but transient acute response. Single-cell transcriptomic analysis of acute LPS-SI unveiled loss of BM monocytes and appearance of an inflammatory monocyte-like (i-Mono’s) population, expressing gene programs similar to early-stage sepsis patients^9^. In the ensuing late phase of LPS-SI, we observed reduced expression of interferon type I (IFN-I) responsive genes in monocytes and profound attenuation of *in vivo* response to a second LPS challenge. Furthermore, late LPS-SI led to impaired myelopoiesis with a loss of intermediate and non-classical monocytes. In accordance, we show compromised myelopoiesis also occurs in late-stage sepsis. Finally, IFNβ treatment reversed LPS-induced immunosuppression in monocytes. Our results reveal long-lasting effects of SI on myelopoiesis and substantiate the importance of IFN-I in the pathophysiology of SI-induced immunosuppression.

## Introduction

SI is a prevalent clinical condition that plays a pivotal role in the pathophysiology of several diseases such as sepsis, which accounts for 20% of all global deaths^1–4^. SI is generally characterized as an initial hyperinflammatory phase followed by a late and prolonged immunosuppressed state^5^. This sustained refractory state of the immune system is associated with high late-onset mortality in sepsis patients^10–12^. Interindividual differences in SI onset time, cause and site of infection, and underlying comorbidities render unraveling molecular mechanisms underlying SI-induced hyperinflammation and immunosuppression in sepsis patients extremely difficult. Animal models of SI are valuable, but suffer from important inter-species differences, thereby limiting translatability^13^. Hence, standardized human SI models capturing hallmarks of both the acute hyperinflammatory and late immunosuppressed phenotypes of sepsis are highly warranted.

Here we utilize human LPS-SI^8^, consisting of intravenous administration of bacterial LPS in healthy volunteers, to elicit transient but profound acute SI followed by a late immunosuppressed state^14^. We employ functional and molecular assays to characterize the immune response to LPS-SI, focusing on the BM compartment. Using single-cell RNA-seq, we reveal that in the acute phase of LPS-SI, an inflammatory monocyte-like population (i-Mono) emerges with gene program similar to that in circulating monocytes of early sepsis patients. Furthermore, we show a clear immunosuppressed phenotype one week after LPS-SI, accompanied by a significant loss of intermediate and non-classical monocytes. We demonstrate that a similar loss of non-classical monocytes occurs in the late phase of sepsis. Importantly, we uncovered type I interferon (IFN-I) signaling impairment in late LPS-SI in the myeloid lineage. Finally, using IFNβ, we were able to restore responsiveness of immunosuppressed monocytes, implicating a pivotal role for impaired IFN-I signaling in sepsis-induced immunosuppression.

## Results

### LPS-SI impact on blood monocytes

To evoke LPS-SI, healthy male volunteers (n=7, **Supp. Table1**) received an intravenous administration of LPS (2ng/kg). The placebo group (n=4) received NaCl 0.9% (**Fig.1a**). All LPS-challenged subjects developed clinical symptoms of SI, such as fever and tachycardia (**Ext. Fig.1a**). Furthermore, LPS-SI was accompanied by a transient increase in circulating levels of pro- and anti-inflammatory cytokines, largely resolved at 8 hours (h) post-LPS (**Ext. Fig.1b**). Severe monocytopenia was observed ∼1 h following LPS administration (**Ext. Fig.1c**). Monocytes started to repopulate the blood ∼3h post-challenge, returning to baseline (d0) levels at ∼6h, virtually all as classical monocytes^15^ (**Ext. Fig.1d**). The following days, monocytes gradually differentiated towards intermediate and non-classical monocytes (**Ext. Fig.1d**)^15^. On day 7 (d7) post-LPS-SI, the abundance of classical monocytes reverted to d0 levels, while numbers of intermediate and non-classical monocytes remained significantly decreased (**Fig.1b**).

**Fig. 1:**
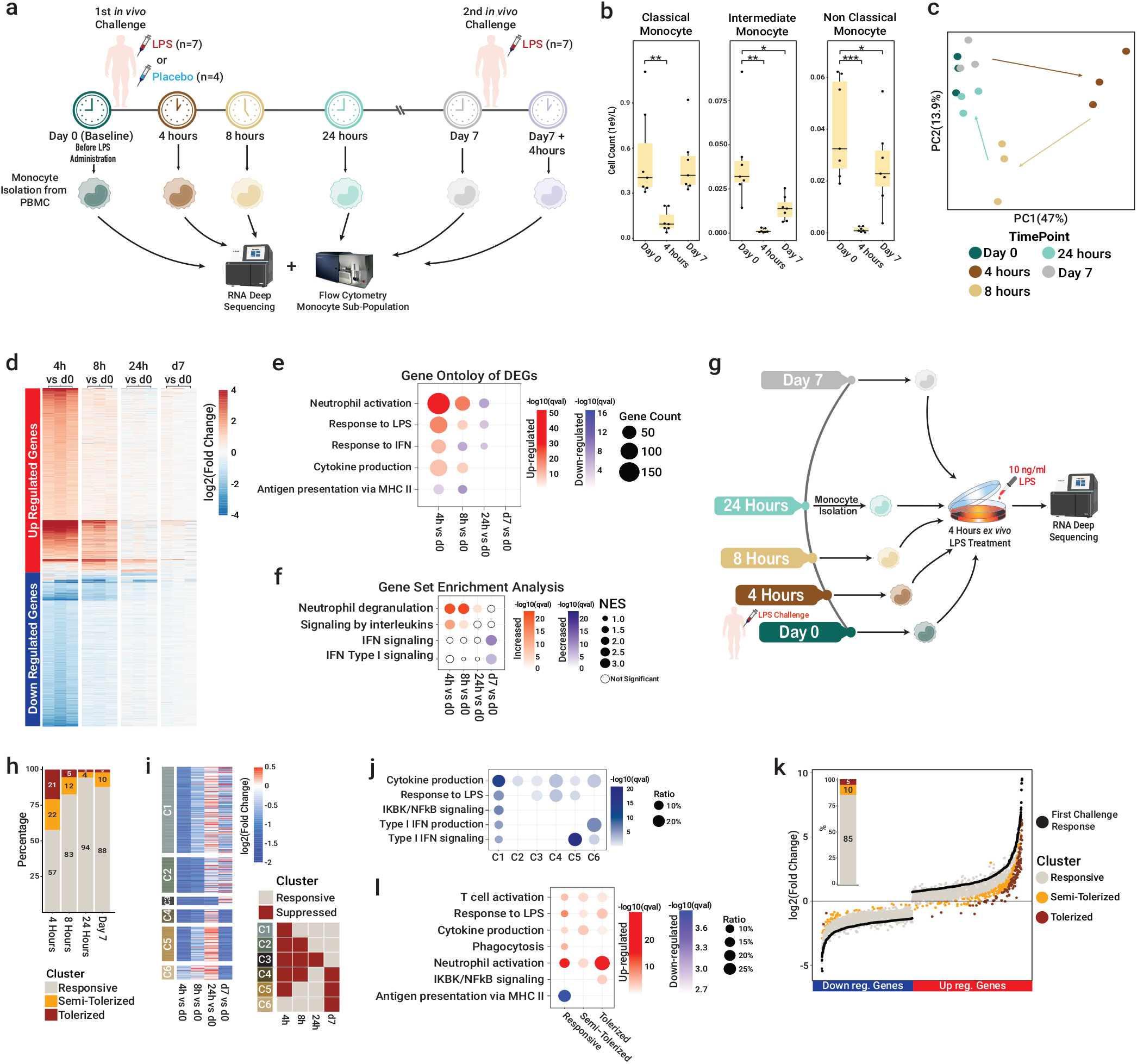
Impairment of IFN-I pathway in blood monocytes one week after LPS-SI. **a**, Schematic representation of the study procedures and blood sample acquisition in healthy volunteers, with seven subjects receiving two *in vivo* LPS challenge (2 ng/kg) with a one-week interval (LPS-induced systemic inflammation [LPS-SI] model) and four subjects receiving placebo (0.9% NaCl). **b**, Absolute abundance of monocyte subtypes in peripheral blood (n=7 for each time point). **c**, Principal component analysis (PCA) of blood monocyte transcriptomes over the LPS-SI time course (n=3 for each time point). **d**, Heatmap representation of differentially expressed genes (DEGs) compared to baseline (d0) over the of LPS-SI time course (n=3 for each time point). **e**, Gene ontology (GO) term analysis of DEGs at each time point. **f**, Gene set enrichment analysis (GSEA) of gene expression profiles at each time point versus d0. **g**, schematic representation of *ex vivo* LPS re-stimulation of monocytes (n=3 for each time point). **h**, Percentage of responsive and (semi-)tolerized genes compared to d0 gene responsiveness. **i**, Heatmap representation of average expression of DEGs based on their relative response to d0 fold-change (n=3). Genes were clustered based on their behavior over the time course. **j**, GO term analysis of genes in each cluster defined in i. **k**, Transcriptomic response of blood monocytes to the second *in vivo* LPS challenge on d7 compared to the response of the first *in vivo* challenge on d0 (n=3). **l**, GO term analysis of (semi-)tolerized genes of monocytes obtained on d7 compared to d0 *in vivo* response. *P* values in (**b**) were calculated using two-sided paired *t*-tests, * = *p* < 0.05; ** = *p* < 0.01; *** = *p* < 0.001.

To characterize monocyte transcriptomic changes during LPS-SI, bulk RNA sequencing (RNA-seq) was performed on isolated circulating CD14+ monocytes obtained from LPS-challenged volunteers (n=3) before LPS administration as well as 4, 8, 24h, and 7 days afterwards (**Fig.1a**). Comparison of gene expression profiles revealed a clear difference at 4 and 8h compared with d0 (**Fig.1c**). Most of the responsive genes returned to d0 expression levels 24h post-LPS with almost no significant differentially expressed gene (DEG) found on d7 (**Fig.1d and Ext. Fig.2a**). Gene ontology (GO) of up-regulated DEGs was mostly attributed to inflammatory response and IFN signaling. Antigen presentation via MHC class-II was the most significant GO term of down-regulated genes at 4h (**Fig.1e and Ext. Fig.2b**). Decreased MHC-II (a.k.a. HLA-DR) expression on monocytes is a hallmark of sepsis-induced immunosuppression and correlates with impaired outcome in patients^16,17^. Intriguingly, gene set enrichment analysis revealed significant down-regulation of IFN signaling on d7 compared with d0 (**Fig.1f**). Accordingly, expression of several interferon-stimulated genes (ISGs) was lower on d7 (**Ext. Fig2c&2d** and **Supp. Table2**).

To assess the effects of LPS-SI on monocyte responsiveness, we performed *ex vivo* stimulation assays. Monocytes isolated from LPS-challenged volunteers (n=3) at d0, 4, 8, 24h, and d7 were *ex vivo* restimulated with LPS for 4h followed by RNA-seq (**Fig.1g**). DEGs were classified into three clusters based on the difference in their responsiveness between d0 and each of the other time points: responsive (Fold-change(FC)<2), semi-tolerized (2<FC<3) and tolerized (FC>3). Approximately 43% of DEGs were (semi-)tolerized at 4h (**Fig.1h** and **Ext. Fig.3a**). This suppressed phenotype was alleviated at 8h and 24h, whereas monocytes showed relatively normal responsiveness. Intriguingly, twelve percent of genes in d7 monocytes were (semi-)tolerized. To explore immunosuppression on the protein level, monocytes obtained on d0 as well as at 4h and d7 were stimulated with 10 different stimuli (i.e., pathogen-associated molecular patterns and heat-killed pathogens). In line with RNA-seq results, a marked attenuation in cytokine secretion was observed at 4h and on d7 compared to d0 (**Ext. Fig.3b**).

Subsequently, we classified dynamic genes from the *ex vivo* restimulated monocytes based on their behavior over time into six clusters (**Fig.1i and Ext. Fig.3c**). GO analysis of each cluster revealed that cytokine production and the inflammatory response were suppressed in almost all clusters (**Fig.1j**). Interestingly, GO terms and individual genes related to type I IFN (IFN-I) signaling were suppressed in LPS-restimulated monocytes obtained on d7 (C5 and C6) (**Fig.1j** and **Ext. Fig.3d**).

These results indicate that one week after LPS-SI, monocytes lack proper expression of IFN-I genes and display impaired induction of ISGs upon *ex vivo* restimulation with LPS and reduced cytokine production capacity upon *ex vivo* stimulation with a range of inflammatory compounds.

### LPS-SI induced *in vivo* immunosuppression

To investigate *in vivo* immunosuppression induced by LPS-SI, LPS-challenged volunteers were re-challenged with the same dose of intravenous LPS administration on d7 (**Fig.1a**). This second challenge resulted in markedly less pronounced clinical symptoms and cytokine elevation compared to the first challenge (**Ext. Fig.3e-g**). This attenuated inflammatory response was also observed in the transcriptome of blood monocytes, with almost all DEGs (93.6%) of the first challenge showing a less pronounced response upon the second (**Ext. Fig.3h**). We clustered DEGs into three groups based on the difference between the first and second challenge: responsive (FC<2), semi-tolerized (2<FC<3) and tolerized (FC>3). This classified 15% of the genes as (semi-)tolerized (**Fig.1k**). GO terms associated with chemokine-mediated response showed suppression, while phagocytosis was still functional on d7 (**Fig.1l**). These results demonstrate that LPS-SI exerts persistent immunosuppressive effects on *in vivo* immune functions. This phenotype is unlikely caused by direct exposition of (pro)monocytes to LPS due to the relatively short life span of monocytes^15^ (∼1-2 days) and their clearance from blood following LPS-SI^15^ along with the rapid clearance of LPS from the circulation (half-life ∼30min)^18,19^.

Collectively, we observed substantially attenuated *in vivo* and *ex vivo* immune responses seven days after LPS-SI. This suggests sustained immunological rewiring of monocytes. It is plausible that the immunosuppressed monocytes originate from stem/progenitor cells in the bone marrow that were exposed to LPS-SI.

### Bone marrow response to LPS-SI

To investigate the effects of LPS-SI on stem/progenitor cells, we determined acute and late changes in transcriptomic profiles of BM-resident cells in LPS-challenged volunteers (n=3). BM samples were obtained before (d0) as well as 4h and 7 days after the first LPS challenge. Mononuclear cells were isolated and single-cell RNA (scRNA-seq) profiles were acquired using the 10X Genomics platform (**Fig.2a**). After quality control and in silico removal of potential doublets, we obtained the transcriptomic profile of 56,506 cells across the three time points (**Supp. Table3**). We visualized cells using uniform manifold approximation and projection (UMAP) embedding, thereby identifying all mononuclear cells from different lineages within the BM (**Fig.2b** and **Ext. Fig.4a**), with relatively similar proportions of cell types between donors (**Ext. Fig.4b**). UMAP visualization at this resolution showed inflammatory monocytes as the most prominent responsive cell type at 4h, followed by inflammatory T cells (**Fig.2c**).

**Fig. 2:**
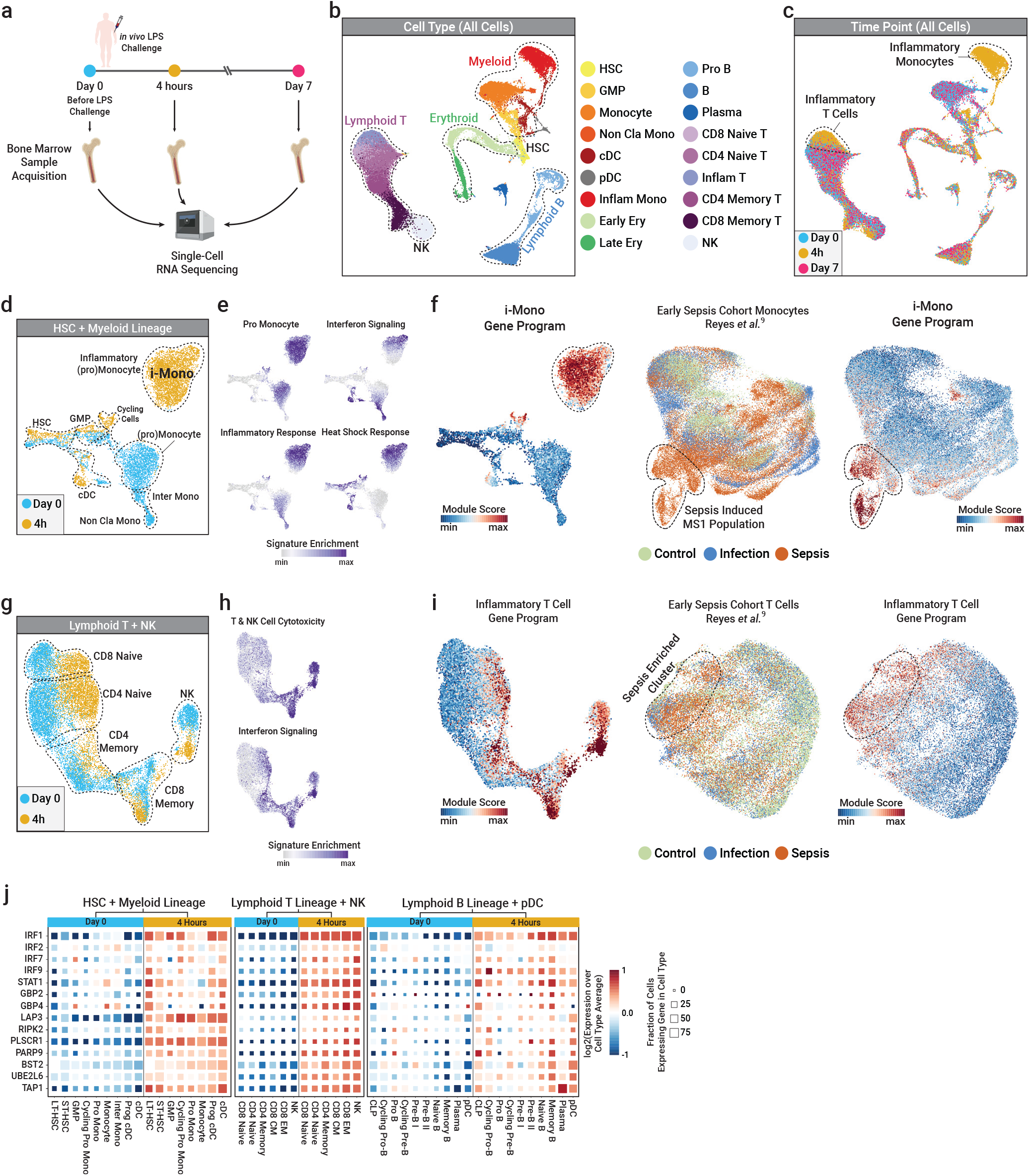
Comprehensive characterization of acute BM response to LPS-SI. **a**, Schematic representation of bone marrow (BM) sample acquisition and subsequent single-cell RNA-sequencing (scRNA-seq) profiling of BM mononuclear immune cells. **b**, Uniform manifold approximation and projection (UMAP) representation of all single cells profiled, colored based on cell type. **c**, UMAP of all cells, colored based on acquisition time point. **d**, UMAP of BM-residing hematopoietic stem (HSCs) and myeloid lineage cells at baseline (d0) and the acute (4h) phase of LPS-SI, colored based on time point. **e**, Single sample gene set enrichment analysis (ssGSEA) of gene sets enriched at 4h myeloid cells compared to d0. **f**, Non-negative matrix factorization (NMF)-inferred gene program enriched in inflammatory monocytes (i-Mono) of LPS-SI (left panel), UMAP of monocytes from a scRNA-seq dataset of an early sepsis patient cohort^9^ (middle panel), enrichment of NMF-inferred i-Mono gene program in early sepsis patient cohort (right panel). **g**, UMAP of BM-residing T and NK cells at d0 and 4h, colored based on time point. **h**, ssGSEA of gene sets higher enriched in 4h T and NK cells compared to d0. **i**, NMF-inferred gene program enriched in 4h T and NK cells (left panel), UMAP representation of T cells from a scRNA-seq dataset of an early sepsis patient cohort^9^ (middle panel), enrichment of NMF-inferred inflammatory T cells gene program in the early sepsis patient cohort (right panel). **j**, Normalized expression profile of several interferon (IFN) pathway genes; square color depicts log2(relative expression at 4h vs. d0), square size is proportional to the percentage of positive (expressing) cells.

We first focused on d0 and 4h to capture the acute phase of LPS-SI. We partitioned cells into 4 lineages: 1) hematopoietic stem cells (HSCs) and myeloid, 2) lymphoid B and plasmocytic dendritic cells (pDCs), 3) lymphoid T and natural killer cells (NKs), and 4) megakaryocyte/erythroid. We did not further analyze the erythroid lineage since differences in RNA profiles at 4h compared to d0 were not evident.

Sub-clustering and batch correction of HSCs and myeloid lineage cells revealed a clear displacement of the entire lineage at 4h, with the most prominent emergence of inflammatory (pro)monocytes at 4h, referred to as i-Monos (**Fig.2d** and **Ext. Fig.4c-d**). To determine molecular pathways underlying the observed cellular variation, we utilized single sample gene set enrichment analysis (ssGSEA). We identified six main gene sets that are enriched along the myeloid lineage (**Ext. Fig.4e**), from which four showed higher enrichment at 4h compared to d0 (**Fig.2e** and **Ext. Fig.4f**). Heat shock response exhibited higher enrichment in all cell types at 4h particularly in the progenitor compartment (**Fig.2e**). Importantly, IFN signaling (both type I and II) also showed enrichment in long-term HSCs as well as in i-Monos and cDCs at 4h (**Fig.2e** and **Ext. Fig.4f**).

Furthermore, we observed a prominent induction of pro-monocyte signature genes in the i-Mono cluster such as RETN, ALOX5AP, and loss of mature monocytes markers such as MHC-II genes (**Ext. Fig.4g**). The higher enrichment of pro-monocyte gene signature was accompanied by an increased proportion of immature pro-monocytes (i-ProMono) at 4h (**Ext. Fig.4h**) This phenomenon is similar to the induction of immature monocytes called myeloid-derived suppressor cells (MDSCs/MS1) observed in sepsis patients^9,20^. Therefore, we sought to compare our BM data to a recently published scRNA-seq dataset of blood samples taken from patients during early bacterial sepsis. We used non-negative matrix factorization (NMF) to define gene programs and determined an i-Mono gene program enriched in BM monocytes at 4h (**Fig.2f**). Projection of this program onto the early sepsis cohort showed a clear enrichment in sepsis-induced monocytes (MS1 population defined by Reyes et al.^9^) (**Fig.2f** and **Ext. Fig.4i**). The reverse approach of projecting the MS1 signature^9^ onto our BM LPS-SI cohort also yielded a clear enrichment of the signature in the i-Mono cluster in BM (**Ext. Fig.4j**).

LPS-induced effects on BM were not restricted to the myeloid lineage. Analysis of lymphoid T and NK cells showed a clear response in all cell types (**Fig.2g** and **Ext. Fig.5a-b**). Interestingly, ssGSEA showed that the IFN signaling signature was one of the causes of displacement on the UMAP (**Fig.2h** and **Ext. Fig.5c-d**). Furthermore, signature sets related to T-NK cell-mediated cytotoxicity were enriched in memory CD8 and NK cells at 4h (**Fig.2h** and **Ext. Fig.5c-d**).

Using NMF, we defined a gene program related to inflammatory T-NK cells that was highly induced at 4h (**Fig.2i**). Projection of this gene program onto the UMAP of circulating T cells obtained from the aforementioned cohort^9^ of early sepsis patients revealed elevated module scores across T cells of patients with severe infection and in particular in those with sepsis (**Fig.2i**). The highest gene module score was observed for the sepsis-enriched cluster (**Fig.2i** and **Ext. Fig.5e**).

Analysis of the lymphoid B lineage and pDCs showed a slight yet notable response of B, plasma cells and pDCs to LPS-SI (**Ext. Fig.5f-h**). SsGSEA revealed 4 major signature sets along the lineage (**Ext. Fig.5i**), with the IFN signaling signature as the enriched gene set in inflammatory memory B and pDCs at 4h (**Ext. Fig.5j**).

Collectively, single-cell RNA-seq analysis of BM mononuclear cells during the acute phase of LPS-SI unveils activation of IFN signaling. Key genes and transcription factors of the IFN pathway are significantly up-regulated particularly in HSCs and myeloid lineage cells as well as in T and NK cells. (**Fig.2j**). Importantly, the comparison of early sepsis patient data with the acute response induced by LPS administration in healthy volunteers revealed extensive similarities, especially pertaining to the induction of immature myeloid cells and to activation of similar molecular gene programs, illustrating its clinical relevance.

### Impaired myelopoiesis and IFN signaling

To explore the sustained immune suppressing effects of LPS-SI on myeloid cells, we included single-cell RNA-seq profiles of HSCs and myeloid lineage cells in BM obtained on d7 post-LPS-SI in our analysis (**Fig.2a**). Cells were analyzed and visualized using UMAP (**Fig.3a**). Interestingly, the transcriptomes of myeloid cells from d7 samples were highly similar compared to those at baseline, except for intermediate and non-classical monocytes (**Fig.3b**). Comparing cell co-embeddings and relative abundance, we observed an approximately two-fold loss of intermediate and non-classical monocytes at d7 compared to d0 (**Fig.3c** and **Ext. Fig.6a**), consistent with our findings in blood (**Fig.1b**). This persistently reduced abundance of intermediate and non-classical monocytes indicates impairment in the monocyte maturation process.

**Fig. 3:**
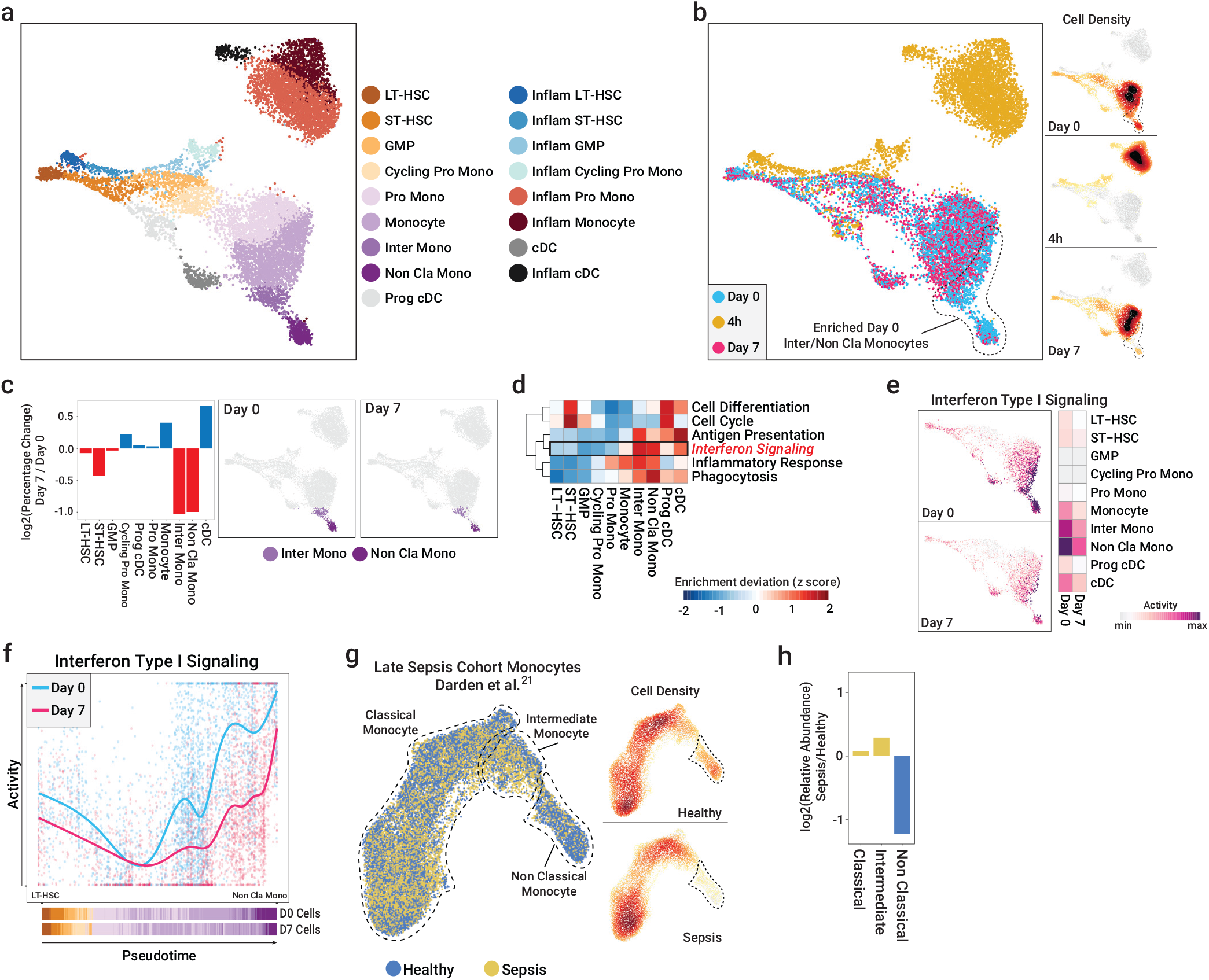
Impairment of myelopoiesis and IFN-I pathway in BM one week after LPS-SI. **a**, UMAP representation of BM-residing HSCs and myeloid lineage cells, colored based on cell type. **b**, UMAP of BM HSCs and myeloid cells, colored based on time point, along with cell density at each BM sample acquisition time point. **c**, log2(relative abundance [percentage] of each cell type) compared between day 7 (d7) and baseline (d0) (left panel), UMAP of intermediate and non-classical monocytes on d0 and d7 (right panel). **d**, Heatmap representation of enrichment deviation (*z*-score) of several signaling pathways for each cell type. **e**, UMAP of IFN-I signaling pathway activity (enrichment) on d0 and d7. Heatmap depicts the average IFN-I activity for each cell type at each time point. **f**, IFN-I activity over the inferred monocyte differentiation trajectory from HSCs to non-classical monocytes, colored based on time point. **g**, UMAP of blood monocytes from a late (>14 days) sepsis patient cohort (left panel^21^), along with cell density calculated for each health status (healthy volunteer or sepsis patient, right panel). **h**, log2(relative abundance) of cell types compared between late sepsis patients and healthy controls.

SsGSEA (**Ext. Fig.6b**) revealed IFN signaling, especially IFN-I, as the most distinguishing signaling pathway of intermediate and non-classical monocytes (**Fig. 3d** and **Ext. Fig.6b**). Projection of IFN-I pathway activity on the UMAP plot demonstrated a clear loss of IFN-I signaling especially in intermediate and non-classical monocytes on d7 (**Fig. 3e** and **Ext. Fig.6c**).

In order to unveil sustained effects of LPS-SI challenge on the entire myeloid lineage, we performed pseudotime differentiation trajectory analysis (**Ext. Fig. 6d**). Assessment of IFN-I enrichment score from LT-HSC to non-classical monocytes reveals clear suppression of this signaling pathway throughout the entire trajectory on d7 (**Fig. 3f**). Furthermore, several key genes and transcriptional factors of IFN-I signaling showed lower expression and fewer positive cells in myeloid lineage on d7 compared to d0, illustrating impaired IFN-I signaling (**Ext. Fig.6e-f**).

To investigate the similarity between sustained effects of LPS-SI and the late, hyporesponsive phase of sepsis, we compared our late BM data to those of a recently published dataset of blood samples from sepsis patients >14 days into the disease^21^. All patients in the cohort had prolonged ICU stays and unresolved organ dysfunction. We extracted the myeloid lineage section of the data and embedded it using UMAP (**Fig.3g** and **Ext. Fig.7a**). Similar to what we found on d7 following LPS-SI, a significant loss of non-classical monocytes was observed in the late-phase sepsis patients (**Fig.3g-h** and **Ext. Fig.7b**). Furthermore, intermediate and non-classical monocytes exhibited the highest IFN-I activity (**Ext. Fig.7c**). However, we did not observe deactivation of IFN-I activity in sepsis patients, which may be due to an ongoing infection, which sustains an ‘active’ inflammatory state in these patients (**Ext. Fig.7c**).

Collectively, our results indicate that LPS-SI resulted in sustained impairment of myelopoiesis, evidenced by lower number of intermediate and non-classical monocytes, and overall reduced IFN-I signaling throughout the myeloid lineage. Furthermore, we observed a similarly impaired myelopoiesis in late-phase sepsis patients.

### Reversal of immunosuppression using IFNβ

Given the impairment of IFN-I signaling as well as the tolerization of IFN-I genes in *ex vivo* LPS-restimulated monocytes (**Fig.1i-j**), we hypothesized that IFNβ reverses LPS-induced immunosuppression. Hence, we performed functional *in vitro* experiments in LPS or mock-treated monocytes exposed to increasing concentrations of IFNβ (**Fig.4a**). IFNβ did not relevantly modulate cytokine responses in non-tolerized cells. However, IFNβ significantly restored production of TNF and IL6 in immunosuppressed cells (**Fig.4b**).

**Fig. 4:**
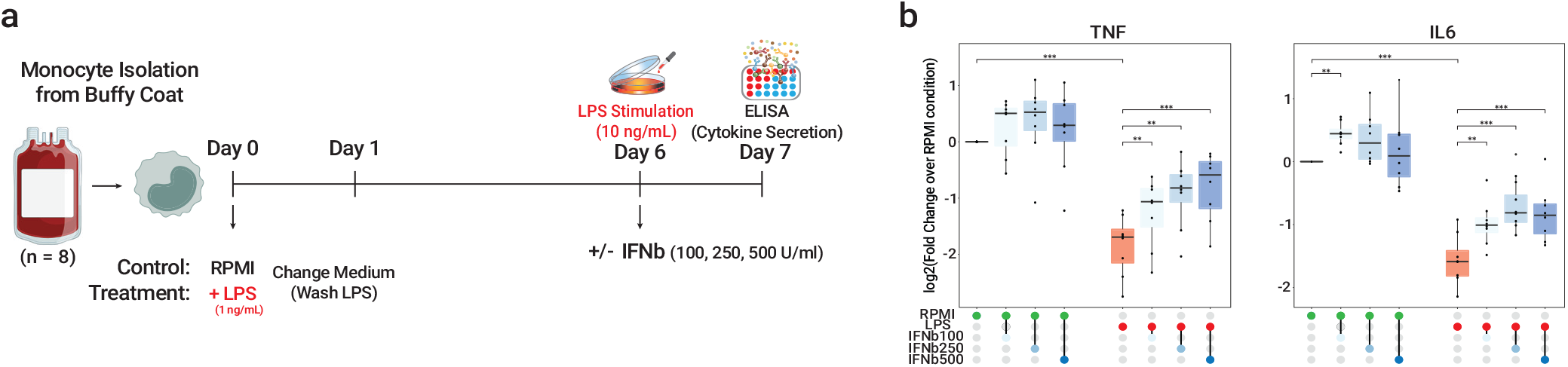
IFNβ treatment reverses LPS-induced immunosuppression. **a**, Schematic representation of reversal experiments with IFNβ in LPS-immuno-suppressed monocytes **b**, log2(fold change) of monocyte cytokine secretion (TNF left panel and IL6 right panel). The fold changes are over RPMI-cultured cells that were stimulated with LPS on day 6 (n=8). *P* values were calculated using two-sided paired *t*-tests, * = *p* < 0.05; ** = *p* < 0.01; *** = *p* < 0.001.

These results indicate that IFNβ reverses the immunosuppressed phenotype of innate immune cells such as monocytes.

## Discussion

Our functional and single cell characterization of LPS-SI revealed the emergence of a specific monocyte-like cell population, termed i-Mono’s, and the temporary loss of normal monocytes during the acute phase of LPS-SI. I-Mono’s express high levels of inflammatory genes and immature promonocyte markers, indicative of emergency myelopoiesis. Strikingly, i-Mono’s have gene programs similar to sepsis-specific MS1 monocytes identified at an early disease stage^9^. Furthermore, we observed activation of IFN signaling in several BM cell types, including HSCs, during the acute phase of LPS-SI. IFN induction in HSCs has been shown to activate dormant HSCs to proliferate and compensate for emergency myelopoiesis, while inducing exhaustion^22–24^.

Previous studies of chronic exposure to pathogens such as Mycobacterium tuberculosis^25^ or metabolites such as heme^26^ in mice showed that HSC modulation can exert long-term effects on myelopoiesis through a process known as ‘innate immune memory’^27,28^.

In our experimental human LPS-SI model, we observed innate immune memory in the form of attenuated *in vivo* responses to a second LPS challenge as well as impaired replenishment of intermediate and non-classical monocytes 7 days after LPS-SI. Furthermore, expression of IFN-I signaling pathway response genes in the myeloid lineage was substantially reduced. It has been reported that mice deficient for either IFNAR1 or IFNβ generate significantly less mature monocytes with reduced expression of ISG^29,30^, suggesting a potential causal link between diminished IFN-I response and impaired myelopoiesis. In accordance, we observed a similar significant loss of non-classical monocytes in late-stage sepsis patients^21^, further substantiating the clinical relevance of the LPS-SI model to other SI-related pathophysiological conditions such as sepsis. Most importantly, our functional experiments revealed that IFNβ reverses the immunosuppressed state of LPS-treated monocytes.

Taken together, impairment of IFN-I signaling appears to play an important role in the development of immunosuppression following SI and, IFNβ may represent a promising treatment option to reverse immunosuppression in SI in general and in sepsis in particular.

## Methods

### Subjects and ethical approval

The study was registered at ClinicalTrials.gov under registration number NCT05570643. This study was approved by the local ethics committee (CMO Arnhem-Nijmegen, The Netherlands, reference no’s NL61136.091.17 and 2017-3337). Eleven healthy male volunteers were recruited. All subjects gave written informed consent and medical history, physical examination, laboratory tests and a 12-leads electrocardiogram did not reveal any abnormalities. Smoking, medication use, previous participation in experimental human endotoxemia, or signs of acute illness within 3 weeks prior to the start of the study were exclusion criteria. All study procedures were performed in accordance with the declaration of Helsinki, including the latest revisions.

### Study design

We performed a randomized placebo-controlled observational study in which subjects were allocated to receive either an intravenous LPS challenge (n=7) or a placebo challenge with 0.9% NaCl (n=4). All LPS-challenged subjects received a second LPS challenge seven days later using identical procedures. Bone marrow and blood were collected at baseline (7 days before first LPS challenge), 4 hours after first endotoxin challenge, and 7 days after the first LPS challenge. Bone marrow aspiration was performed by a skilled physician assistant of the hematology department at Radboudumc. Bone marrow was collected in a sodium heparin solution (150 IE/mL, ratio 3:1). Blood was collected in tubes containing ethylenediaminetetraacetic acid (EDTA) as anticoagulant. Additional blood was collected before and at several timepoints following the challenges.

### Experimental human LPS-SI

During LPS/placebo challenge days, all subjects underwent the same study procedures, except for administration of either LPS or placebo. Briefly, 24 hours before hospitalization, subjects needed to refrain from alcohol and caffeine and from 22:00 onwards no food and drinks were allowed. Prior to the challenge, subjects were admitted to the intensive care research unit of Radboudumc in Nijmegen. An intravenous cannula was placed in an antebrachial vein to administrate fluids and LPS or 0.9% NaCl. A radial artery catheter was inserted to withdraw blood and monitor blood pressure continuously. Prehydration (1.5L 2.5% Glucose/0.45% NaCl) was administrated intravenously in the hour prior to the challenge. Thereafter, a bolus of 2 ng/kg LPS (*E. Coli Type O113*, Lot no. 94332B1; List Biological Laboratories) or saline (placebo) was administered intravenously and hydration fluid (2.5% Glucose/0.45% NaCl) was continued at an infusion rate of 150 mL/h for 8 hours. During hospitalization, heart rate was monitored using a 4-lead electrocardiogram (M50 Monitor, Philips). Every 30 minutes, core temperature was measured with a tympanic thermometer (FirstTemp Genius 2, Covidien) and LPS-induced symptoms (headache, nausea, cold shivers, muscle- and back pain) were scored using a numeric six-point scale (0 = no symptoms, 5 = worst symptoms experienced ever) with addition of 3 points in case of vomiting, resulting in a total symptom score ranging from 0 to 28.

### Cell counts and plasma cytokine measurements

Blood cell counts were analysed using a Sysmex XE-5000 (Sysmex). For cytokine determination, blood was centrifuged directly after withdrawal (10min, 2400g, 4^°^C) and plasma was stored at -80^°^C until analysis. Concentrations of TNF, IL-6, CXCL8 (IL-8), IL-10, CCL3 (MIP-1α), CCL2 (MCP-1), CCL4 (MIP-1β), IL-1RN (IL-1RA) were determined in one batch using a simultaneous luminex assay (Milliplex, Millipore) on a MagPix instrument (Luminex).

### Flowcytometry of monocyte subtypes

Blood was phenotyped with antibodies against CD45-Cy5.5 (A62835, Beckman Coulter), CD14-ECD(B92391, Beckman Coulter), CD16-PE (332779, BD Biosciences), CD64-FITC (B49185, Beckman Coulter), CD11b-PC7 (A54822, Beckman Coulter), HLA-DR-APC (IM3635, Beckman Coulter), DRAQ7 (DR71000, Biostatus), CD192-BV421 (564067, BD Biosciences), CD15-KO (B01176, Beckman Coulter), and dumpgate: CD3-AA750 (A94680, Beckman Coulter), CD19-APCA750 (A94681, Beckman Coulter) and CD56-APC A750 (B46024, Beckman Coulter) on a Navios flow cytometer (Beckman Coulter) at the Hematology department of Radboudumc. Monocyte subtype populations were determined using the gating strategy depicted in **Ext. Fig.8a**.

### *Ex vivo* stimulation of CD14+ monocytes

Peripheral blood mononuclear cells (PBMCs) were isolated using Ficoll-based density gradient separation (1200g, 10 minutes, room temperature, with brake) in SepMate™-50 tubes (Stemcell Technologies). Cells were washed with cold PBS (1700 rpm, 10 minutes, 4^°^C), resuspended in culture medium (RPMI 1640, Gibco) and counted using a Sysmex XE-5000 (Sysmex Nederland). All samples were kept on ice in between procedures. PBMCs were subsequently depleted from neutrophils and intermediate and non-classical monocytes using CD16 microbeads (Miltenyi Biotec) according to the manufacturers protocol. Hereafter, classical monocytes were positively selected using CD14 microbeads (Miltenyi Biotec). Cells were resuspended in culture medium at a concentration of 1*10^6^ cells/mL, and 1*10^5^ cells were seeded in flat-bottom 96-well plates. The cells were incubated with culture medium or various stimuli (Pam3Cys [10 µg/mL, InvivoGen], Poly:IC [50 µg/mL; InvivoGen], *E. coli* LPS [10 ng/mL, serotype 055:B5; Sigma-Aldrich], flagellin [10 µg/mL, InvivoGen], resiquimod [R848, 0.35 µg/mL, InvivoGen], heat killed *E. coli* [10^7^ per well, ATCC35218], *S. aureus* [10^7^ per well, ATCC25923], *P. aeruginosa* [10^7^ per well, PA01], *C. albicans* [10^6^ per well, UC820], *A. fumigatus* [10^7^ per well (with 10% human pooled serum), V05-27]) for 24 hours at 37^°^C with 5% CO_2_. Hereafter, supernatants were collected and stored at -80^°^C until analysis. The concentrations of TNF, IL-1β, IL-1RN (IL-1RA), IL-6, IL-10, CCL4 (MIP-1β) in supernatants of *ex vivo* stimulated cell cultures were determined in one batch using a simultaneous Luminex assay (Milliplex, Millipore) on a MagPix instrument (Luminex).

For gene expression profiling of monocytes after 4 hours of *ex vivo* LPS (re)stimulation, monocytes were isolated from LPS-SI volunteers at 0, 4, 8, and 24 hours as well as 7 days following *in vivo* LPS administration. PBMCs were obtained as described above and monocytes were isolated by positive selection using CD14 microbeads (Miltenyi Biotec). Cells were resuspended in culture medium (RPMI 1640) supplemented with 10% human serum (Sigma Aldrich) and seeded in flat-bottom 96 well plates (2⋆10^5^ cells per well). Cells were left to attach for one hour at 37^°^C with 5% CO_2_, after which culture medium was refreshed, and monocytes were (re)stimulated with 10 ng/mL LPS (Sigma Aldrich) for 4 hours at 37^°^C with 5% CO_2_. After stimulation, monocytes were lyzed using Trizol (Invitrogen) and stored at -80^°^C until further processing.

### Reversal of immune suppression using IFNβ

PBMCs where isolated from blood of 8 healthy volunteers as described in the previous paragraph. Subsequently, monocytes were isolated using Percoll (Sigma Aldrich) density gradient centrifugation (580g, 15 minutes [no brake and slow acceleration], room temperature). Cells were washed with cold PBS (350g, 7 minutes, 4^°^C) after which they were resuspended to a concentration of 1*10^6^/ mL in RPMI. Thereafter, 1*10^5^ monocytes were seeded in flat-bottom 96-well plates and left to attach for one hour at 37^°^C with 5% CO_2_. Non-adherent cells were subsequently removed by washing with warm PBS. Adherent cells were incubated for 24 hours with either culture medium (RPMI) or 1 ng/mL LPS at 37^°^C with 5% CO_2_. After 24 hours, cells were washed with warm PBS. Thereafter, cells were incubated with RPMI supplemented with 10% human pooled serum for 5 days and culture medium was refreshed on day 3. On day 6, culture medium was removed and cells were incubated for 24 hours with LPS (10 ng/mL, serotype 055:B5; Sigma-Aldrich] in the presence and absence of IFNβ (100, 250, and 500 U/mL, R&D systems) for 24 hrs at 37^°^C with 5% CO_2_. Afterwards, supernatants were collected and stored at -80^°^C until determination of TNF and IL-6 using ELISA (R&D systems).

### Total RNA extraction and cDNA synthesis

Total RNA was extracted from lyzed monocytes (isolated as described above) using the RNeasy RNA extraction kit (Qiagen), incorporating on-column DNaseI (Qiagen) DNA digestion. Afterwards, ribosomal RNA was removed using riboZero rRNA removal kit (Illumina). The efficiency of rRNA removal was confirmed using quantitative RT-PCR with primers for GAPDH (as internal control) and 18S and 28S rRNA. RNA molecules fragmented into ∼200bp fragments by incubating in fragmentation buffer (200mM Tris-Acetate, 500mM Potassium Acetate, 150mM Magnesium Acetate [pH 8.2]) for 7.5 min at 95^°^C. First strand cDNA from fragmented RNA was synthesized using SuperScript III reverse transcriptase enzyme (Life Technologies) according to the manufacturer’s protocol and followed by second strand cDNA synthesis.

### Bulk RNA-seq library preparation and sequencing

Gene expression libraries were prepared using KAPA HyperPrep kit (KAPA Biosystems) according to the manufacturer’s protocol. In brief, synthesized double stranded cDNA was incubated with end repair and A-tailing buffer and enzyme initially for 30 min at 20^°^C and then for 30 min at 65^°^C. Library-specific adapters were ligated to tailed DNA molecules using DNA ligase enzyme by incubating for 15 min at 15^°^C. Ligation reaction was cleaned up using Agencourt AMPure XP reagent (Beckman Coulter) and subsequently amplified using 10 cycles of PCR. Finally, 300bp fragments were selected using 2% E-gel selection system (Invitrogen). Size selection was validated with 2100 BioAnalyzer system (Agilent). Prepared libraries were sequenced utilizing NextSeq 500 machine (Illumina) with a paired-end sequencing setup.

### Bulk RNA-seq data analysis

Raw RNA-seq reads were aligned to hg38 reference genome and gene expression profiles were quantified using STAR aligner^31^. Genes with less than 50 mapped reads on condition average were excluded from the analysis. For each comparison of RNA-seq profiles, gene expression data of corresponding samples was normalized and differentially expressed genes were identified using DESeq2^32^ analysis package utilizing fold-change >2 and q value (Benjamini-Hochberg adjusted p value) <0.05 as statistical significance cutoffs.

### Gene Ontology and Gene Set Enrichment Analysis of bulk RNA-seq data

To infer significantly enriched gene ontologies for identified gene sets of interest, such as differentially expressed gene, we used clusterProfiler^33^ analysis package. Gene set enrichment analysis (GSEA) for the comparison of day 0 and day 7 monocyte gene expression profiles was done using fgsea^34^ package.

### Single-cell RNA-seq sample preparation and sequencing

For single-cell RNA-seq (scRNA-seq), mononuclear cells were isolated from bone marrow samples using identical procedure as used for PBMCs (see above) and cryopreserved at -80^°^C until further processing. Single Cell Gene Expression 3’ v3 (10X Genomics) was utilized following the manufacturer’s protocol. In brief, approximately 10,000 to 15,000 single cells were loaded into a channel of Chromium chip and loaded chip was inserted into the Chromium Controller. After generation of single-cell gel bead-in-emulsion (GEMs) and reverse transcription of RNA, cDNA amplification, fragmentation and adapter ligation were done. The quality of prepared sequencing libraries was assessed using 2100 Bioanalyzer (Agilent). scRNA-seq libraries were sequenced on a NextSeq500 (Illumina) or NovaSeq (Illumina).

### Single-cell RNA-seq data preprocessing and analysis

Demultiplexing the raw BCL files was done using Cell Ranger mkfastq (v3.1.0) software and resulting fastq files were mapped to human GRCh38 reference genome using Cell Ranger count software with default parameters. Output count matrix was imported to R analysis software and further analyzed using Seurat (v4.0.4)^35^. Low quality cells with mitochondrial percentage above 15% or with fewer than 200 genes and fewer than 40,000 UMI counts were excluded from the analysis. Cell expressing multi canonical lineage markers at the same time, such as T and B cells specific markers, were identified as potential doublets and removed from the analysis. Afterwards, gene expression profiles were normalized to sequencing depth and scaled to 10,000 counts and log transformed. Batch correction for inter-donor differences was done utilizing RunFastMNN function from SeuratWrappers package which is the R implementation of MNN^36^ batch correction method. Cells were embedded on 2D view using UMAP and clustered using FindClusters function from Seurat package with the resolution of 1. Cluster/Cell type specific marker genes were identified using FindMarkers function from Seurat package with the default parameters. Based on well-known cell type markers we annotated each cluster and visualized violin plot of gene expression for several cell type specific markers using stacked_violin function of scanpy^37^ analysis package in Python (v3.7.7). Cell Density plots are generated using embedding_density function of scanpy package. For lineage-specific analysis the same steps were done by initial extract of corresponding lineage cells from the whole bone marrow dataset and further embedding and clustering. Unless specified all figures were generated using ggplot2 visualization package and all heatmaps were generated using pheatmap package in R (v4.1.0).

### Pseudotime trajectory analysis of scRNA-seq data

In order to identify underlying differentiation trajectory starting from long-term HSCs to mature non-classical monocytes we performed pseudotime trajectory analysis using monocle3^38^ analysis package. We utilized the suggested workflow of the monocle3 with minimal_branch_len parameter set to 5 and rann.k parameter set to 30. For each time point obtained pseudotime values were normalized to [0-1] range with the most immature stem cell having the 0 value and the most mature non-classical monocyte having 1 value. Cells were visualized using ggplot2 package.

### Single-sample GSEA (ssGSEA) and Non-Negative Matric Factorization (NMF)

To identify signaling pathways responsible for variations observed between cell types and time points we performed ssGSEA using VISION^39^ analysis package. We utilized hallmark, gene ontology and Reactome gene sets from molecular Signature Database (MSigDB)^40^. After calculation of the enrichment of each gene set for each single cell, in order to cluster similar gene sets into one meta gene set, we measured the Pearson correlation of different gene sets and clustered highly similar gene sets into one meta gene set, which we defined as signature set. Using terms in each signature set we annotated each of them. We performed same analysis for each of the three lineages studied in this article (HSC + myeloid, B + pDC and T +NK lineages).

To identify the underlying gene program responsible for the generation of i-Monos and inflammatory T cells at 4h time point of LPS-SI experiments we performed NMF using RunNMF function of STutility^41^ package with the default parameters obtaining 40 factors/gene programs. Afterwards, highly similar gene programs were clustered together generating a meta gene program. The identified meta gene program highly enriched in i-Monos and inflammatory T cells were visualized using ggplot2. Top genes contributing to each of gene programs are listed in **Supp. Table 4**.

### Analysis of early and late sepsis patient data

For the comparison of results obtained from LPS-SI study with data obtained from patients in early and late phase of sepsis, we downloaded publicly available scRNA-seq datasets. For the early sepsis dataset, we downloaded data from the Broad Institute Single Cell Portal with the accession number of SCP548 and extracted monocytic and T cell compartments of the data using cell annotations from the corresponding dataset. As recommended in the published article^9^ we also did not perform any batch correction on the obtained data and embedded and visualized cells using UMAP. The MS1 gene set acquired from the corresponding publication^9^.

For the late sepsis dataset, we downloaded data from Gene Expression Omnibus (GEO) database with the accession number of GSE175453 and extracted monocytic compartment for the dataset using the cell annotations from the corresponding dataset. We performed batch correction for inter-donor differences using RPCA function from Seurat package and visualized cells using UMAP. We generated cell density plot for the healthy and late sepsis samples using the embedding_density function of scanpy package.

## Data Availability

Bulk RNA-seq and scRNA-seq data for this study can be downloaded from GEO database (https://www.ncbi.nlm.nih.gov/geo/) with the accession number: GSE212093.

## Code availability

Codes used to perform the data analysis related to this study are available at https://github.com/fkeramati/LPS-SI

## Acknowledgments

F.K. is supported by the Princess Maxima Center and ZonMW-TOP project 91-21-6061. C.R.M. is supported by the Princess Maxima Center and the European Union’s Horizon 2020 Skłodowska-Curie Actions (project AiPBAND) under grant #764281. W.M. is supported by the Italian National Operational Programme on Research 2014-2020 (PON AIM 1859703-2). H.G.S. is supported by the Princess Maxima Center and Kika (Kinderen Kankervrij).

## Author contributions

F.K., G.P.L., H.G.S., P.P. and M.K. conceived and designed the experiments with contributions from N.B., I.G., E.v.G., K.T., S.vS., W.v.d.V., F.P., J.G. and M.G.N. F.K., G.P.L., N.B., A.PvT., H.H., M.B., E.v.G., K.T., J.G. performed experiments related to *in vivo* LPS challenge and *ex vivo* stimulations. F.K. and G.P.L. analyzed data with contributions from N.B. and W.M. F.K. prepared single cell libraries with contributions from C.R.M. F.K. analyzed and interpreted single cell data with contributions from E.H. F.K., G.P.L., H.G.S. and M.K. wrote the manuscript. H.G.S and M.K. supervised the study. All authors critically reviewed and approved the manuscript.

---

**Extended Fig. 1:**
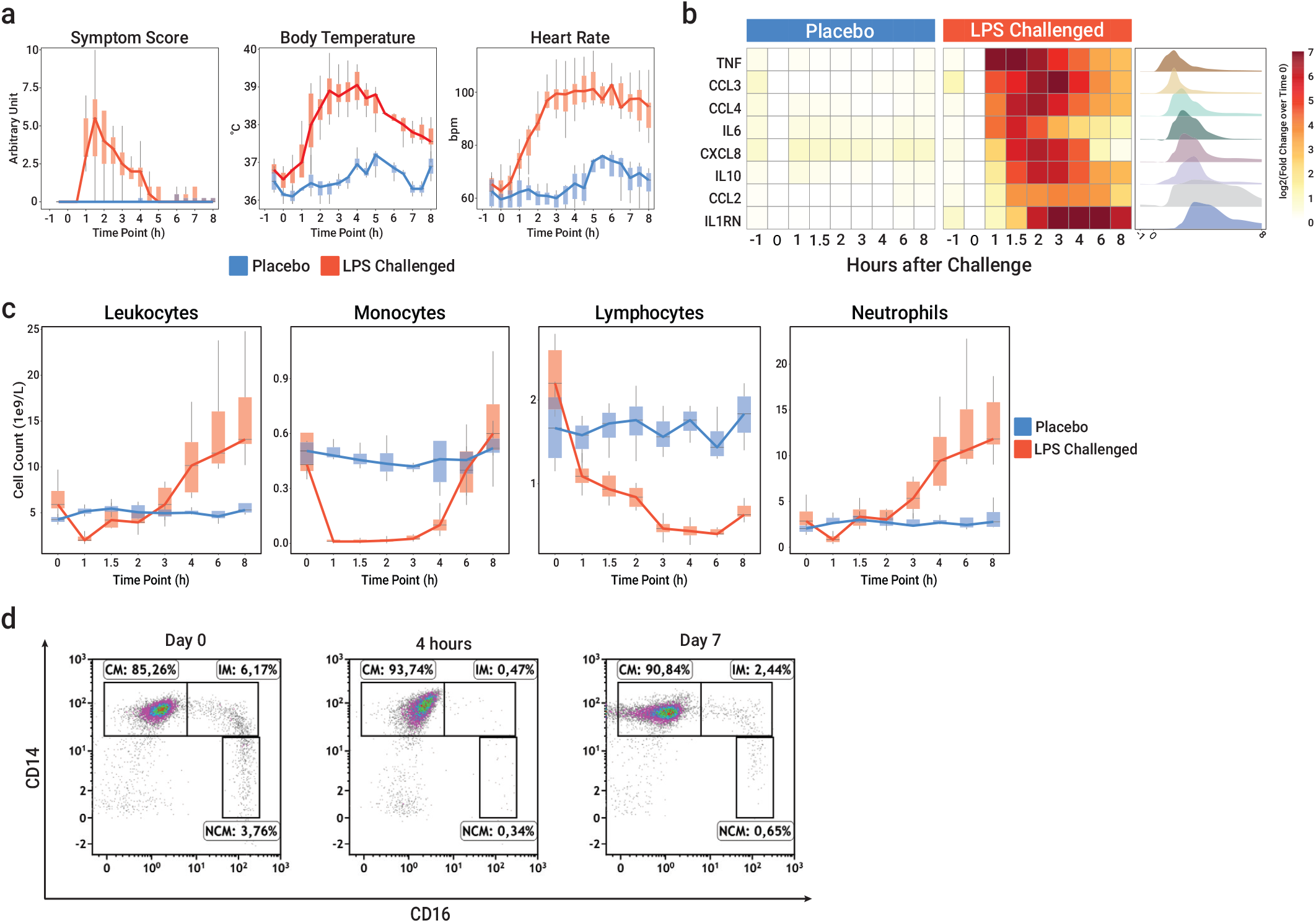
Phenotypical characteristics of LPS-SI. **a**, Vital sign changes upon an *in vivo* challenge with LPS (red, n=7) or placebo (blue, n=4). **b**, Profile of circulating inflammatory cytokines in blood of LPS- (n=7) or placebo-challenged (n=4) individuals. **c**, Absolute number of leukocytes, monocytes, lymphocytes and neutrophils in blood of LPS (n=7) or placebo-challenged (n=4) individuals **d**, Representative example of flow cytometric analysis of different monocyte subsets before (d0) and various timepoints following LPS administration. Boxplots in (**a** and **c**) represent data distribution within first and third quartiles, with whiskers indicating range.

**Extended Fig. 2:**
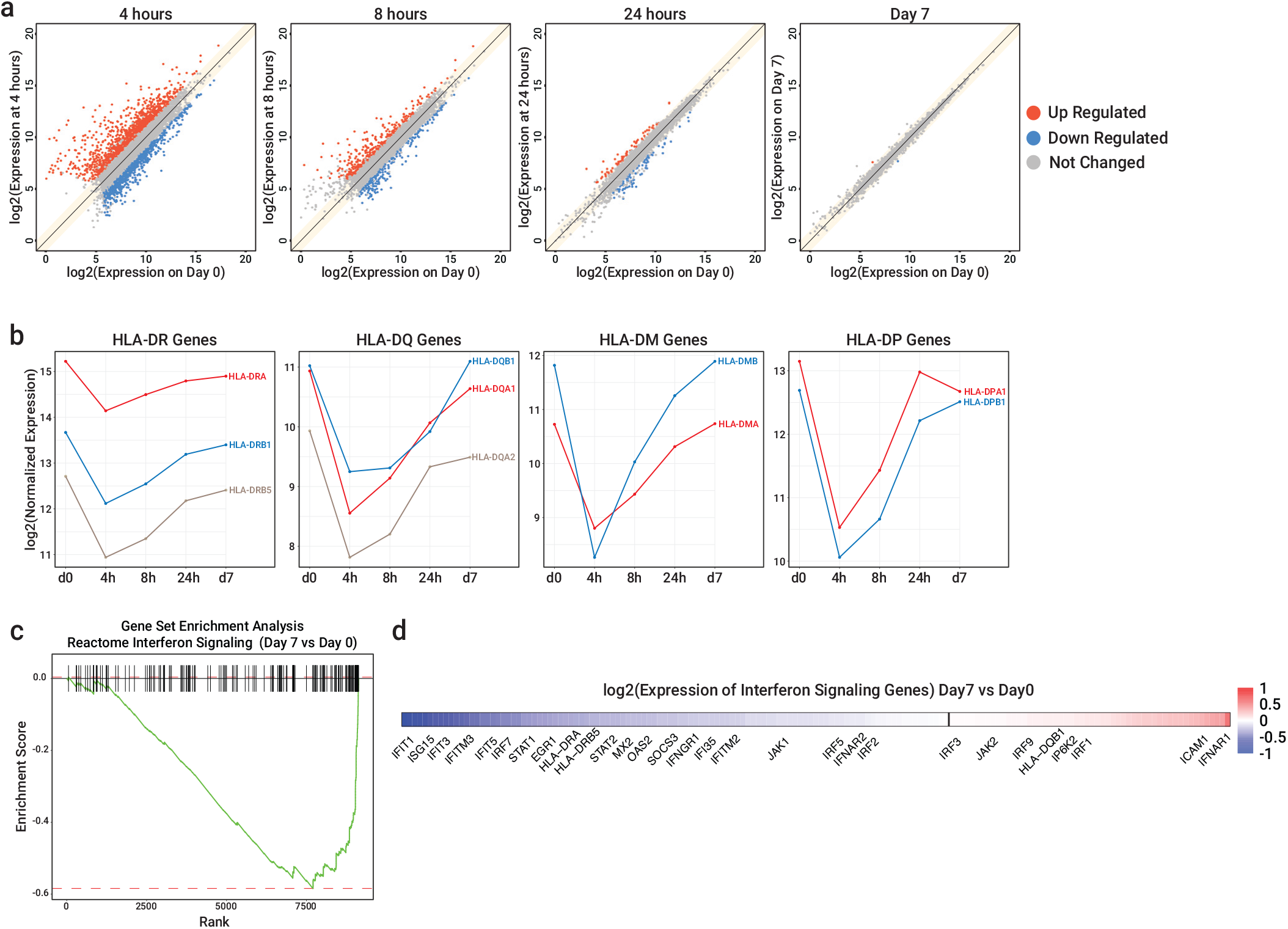
IFN signaling impairment in blood monocytes one week after LPS-SI. **a**, Scatterplot of the average gene expression at different timepoints following LPS administration (y-axis) versus baseline (d0, before LPS administration, x-axis). Dots represent the average of expression of n=3 replicates. Red dots indicate significantly up-regulated genes (fold-change > 2 & *q* value < 0.05), while blue dots depict significantly down-regulated genes (fold-change < 0.5 & *q* value < 0.05). **b**, log2(mean normalized expression) of MHC class II (HLA-II) genes (n=3) at different timepoints following LPS administration. **c**, Gene set enrichment analysis of interferon signaling genes in peripheral blood monocytes (based on the comparison of d7 vs. d0). **d**, log2(relative normalized expression) of interferon signaling genes, ordered and colored based on relative expression on d7 vs. d0.

**Extended Fig. 3:**
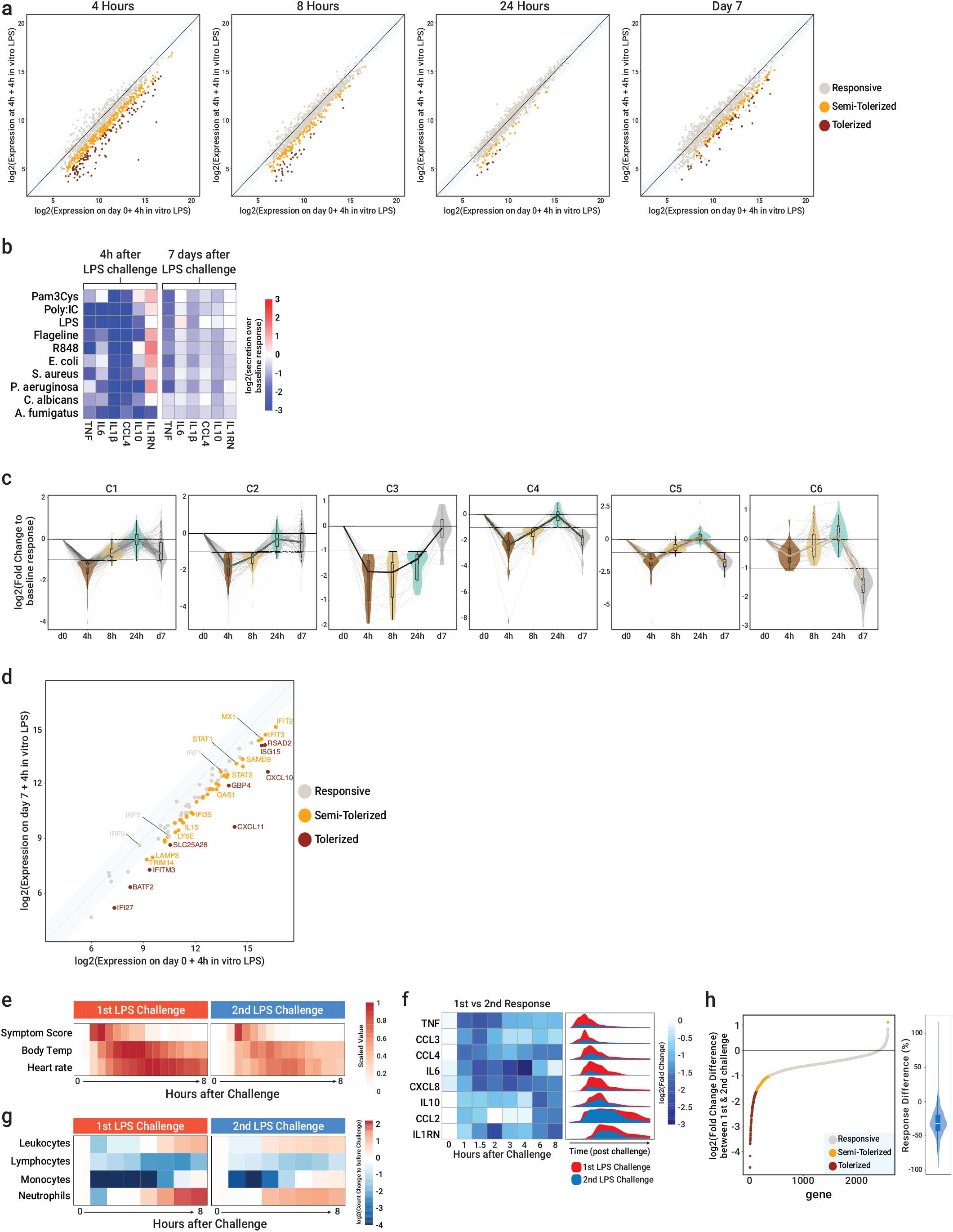
Immune suppression one week after LPS-SI. **a**, Scatterplot of mean normalized gene expression profiles of monocytes (n=3) obtained at various timepoints following LPS administration that were stimulated *ex vivo* with LPS for 4h. For each timepoint the log2(normalized expression) of LPS stimulated monocytes at that timepoint (y axis) is compared to log2(normalized expression) of LPS-stimulated monocytes at baseline (d0, x axis). Orange dots indicate semi-tolerized genes (−1>log2[fold-change]>-2) and red dots indicate tolerized genes (−2>log2[fold-change]). **b**, Heatmap of average log2(fold-change) difference in cytokine production by monocytes obtained in the acute (4 hours post-LPS challenge) and late phase (7 days post-LPS challenge) that were *ex vivo* stimulated with various stimuli; comparison with monocytes obtained at baseline (d0) that were *ex vivo* stimulated with the same stimuli (n=7). **c**, violin plot of log2(fold-change) difference between *ex vivo* LPS-stimulated monocytes obtained at various timepoints following LPS administration and LPS-stimulated monocytes obtained at baseline (d0). Genes are clustered based on their behavior over time (see main text and **Fig. 1i**). **d**, Scatterplot of log2(normalized expression) of IFN-I signaling pathway genes in monocytes that were isolated from blood of LPS-SI individuals at d7 and *ex vivo* re-stimulated with LPS (y-axis, mean expression of n=3) compared to d0 monocytes *ex vivo* stimulated with LPS (x-axis, mean expression of n=3). **e**, Comparison of changes in vital sign changes during the first and second LPS challenge (n=7). **f**, log2(fold-change) difference between plasma cytokine levels during the first and second LPS challenge (n=7). **g**, Comparison of leukocyte (subtypes) during the first and second LPS challenge (n=7). **h**, log2(fold change) difference between monocytic transcriptomic response to the first and second LPS challenge (n=3).

**Extended Fig. 4:**
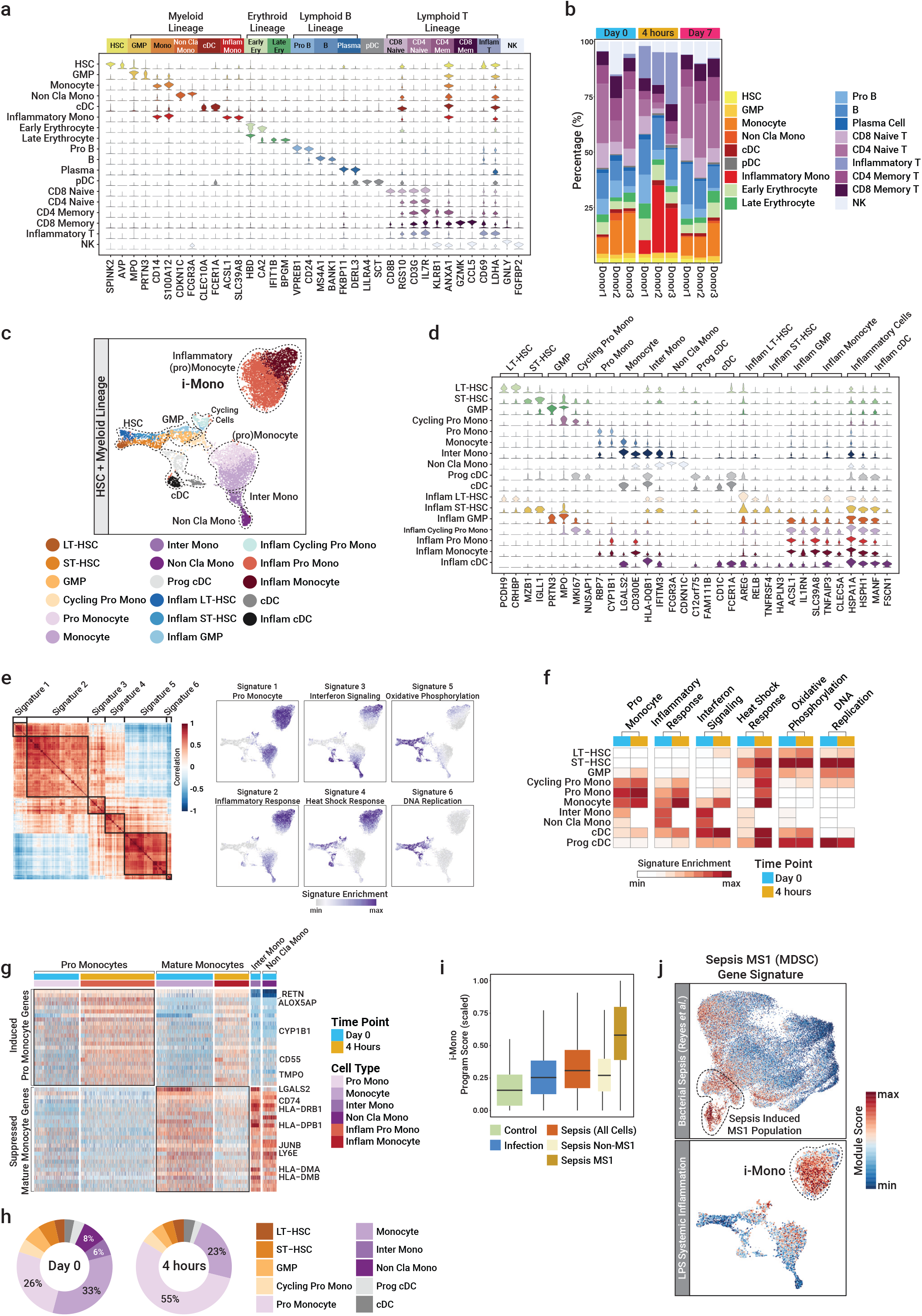
Single-cell transcriptome analysis of acute response to LPS-SI in BM-residing cells. **a**, Marker genes of all mononuclear immune cell types identified in BM. **b**, BM composition (percentage) for each cell type depicted for each donor and timepoint. **c**, UMAP of HSCs and myeloid lineage cells obtained from baseline (d0) and 4h post-LPS BM samples (n=3) colored by cell type. **d**, Marker genes of HSCs and myeloid cell types identified in BM. **e**, Correlation heatmap of 6 major signature gene sets determined using ssGSEA (left panel), UMAP representation of identified 6 gene sets with relative enrichment on single cell level (right smaller panels). **f**, Average enrichment of 6 identified gene sets per cell type and timepoint. **g**, Heatmap representation of promonocyte and mature monocyte genes in myeloid cells. Each column represents a single cell and each row represent a gene, colored based on normalized gene expression (red indicates higher expression). **h**, Percentage of each cell type at each time point (d0 and 4h post-LPS). **i**, Distribution of inflammatory monocyte (i-Mono) gene program enrichment for each condition (healthy, infection, sepsis) in the early sepsis patient cohort. **j**, Enrichment of sepsis specific MS1 (MDSC) gene signature defined in early sepsis patients by Reyes et. al^9^ (upper panel), and LPS-SI BM myeloid cells (lower panel).

**Extended Fig. 5:**
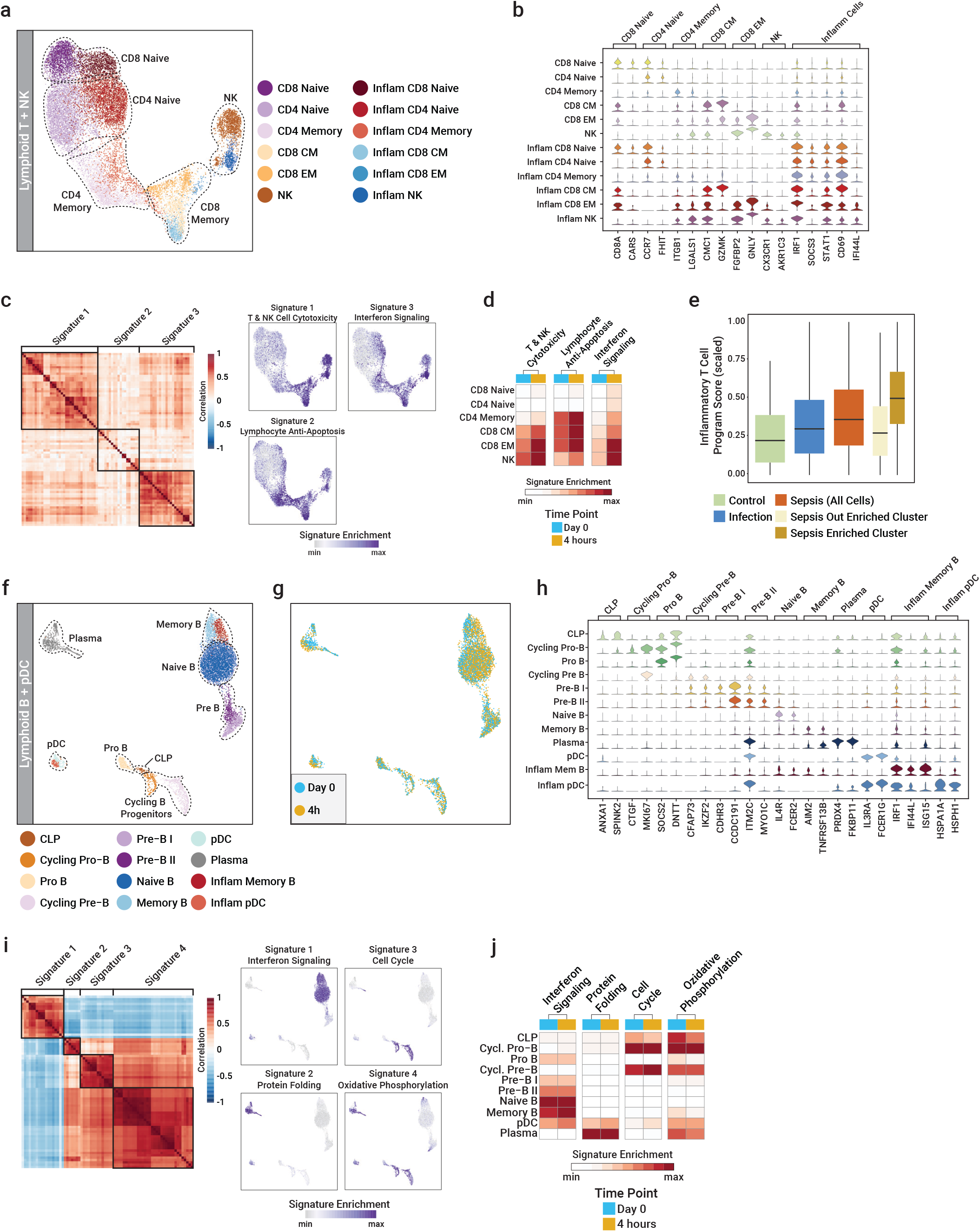
Single-cell transcriptomic analysis of acute responses to LPS-SI in BM-residing lymphoid T and B and NK cells. **a**, UMAP of T and NK cells obtained from baseline (d0) and 4h post-LPS BM samples (n=3) colored by cell type. **b**, Marker genes of different T and NK cell types identified in BM. **c**, Correlation heatmap of 3 major signature gene sets determined using ssGSEA (left panel), UMAP representation of identified 3 gene sets with relative enrichment on single cell level (right smaller panels). **d**, Average enrichment of 3 identified gene sets per cell type and time point. **e**, Distribution of inflammatory T cells gene program enrichment for each condition (healthy, infection, sepsis) in the early sepsis patient cohort. **f**, UMAP of B lineage cells and pDCs obtained from baseline (d0) and 4h BM samples (n=3) colored by cell type. **g**, UMAP of B lineage and pDCs, colored based on BM acquisition time point. **h**, Marker genes of different B lineage and pDCs identified in BM. **i**, Correlation heatmap of 4 major signature gene sets determined using ssGSEA (left panel), UMAP representation of identified 4 gene sets with relative enrichment on single cell level (right smaller panels). **j**, Average enrichment of 4 identified gene sets per cell type and timepoint.

**Extended Fig. 6:**
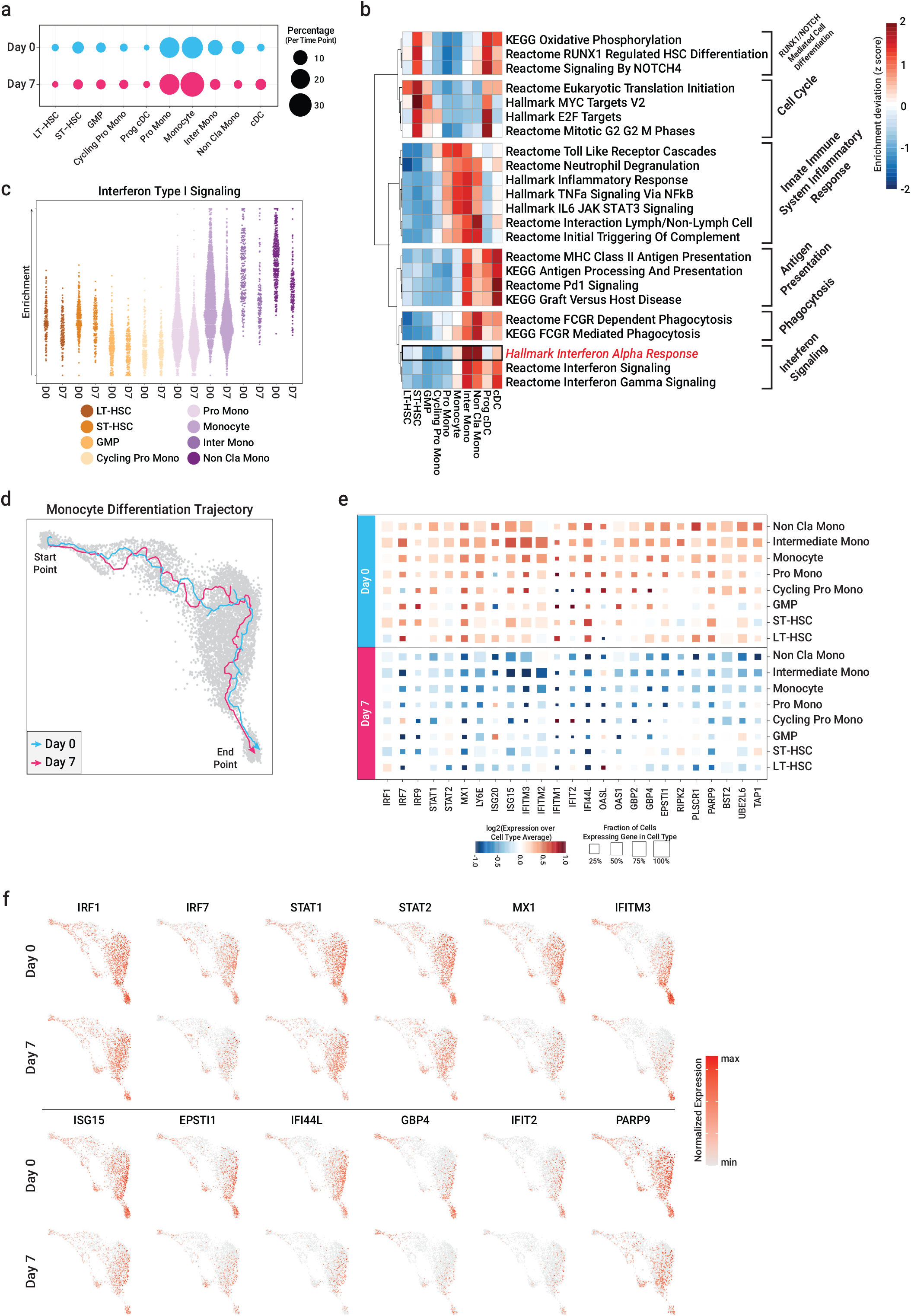
Impairment of IFN-I one week following LPS-SI in BM myeloid lineage cells. **a**, Cell type proportion (percentage) in BM-residing HSC and myeloid lineage cells per timepoint. **b**, Heatmap representation of enrichment deviation (z-score) of several signaling pathways for each cell type, with the most differentially enriched pathway for each cell type colored in red. Signaling pathways are clustered into 6 major pathways based on their enrichment profile for different cell types. **c**, IFN-I enrichment (activity) for single cells grouped per cell type and time point (d0 and d7). **d**, Monocyte differentiation pseudotime trajectory starting from LT-HSC (start point) to non-classical monocytes (end point) colored by time point. **e**, Normalized expression profile of several interferon (IFN) pathway genes; square color depicts log2(relative expression at d7 vs. d0), square size is proportional to the percentage of positive (expressing) cells. **f**, UMAP representation of normalized single-cell gene expression of several IFN-I signaling pathway genes on d0 and d7. Red dots represent positive gene expressing single cells.

**Extended Fig. 7:**
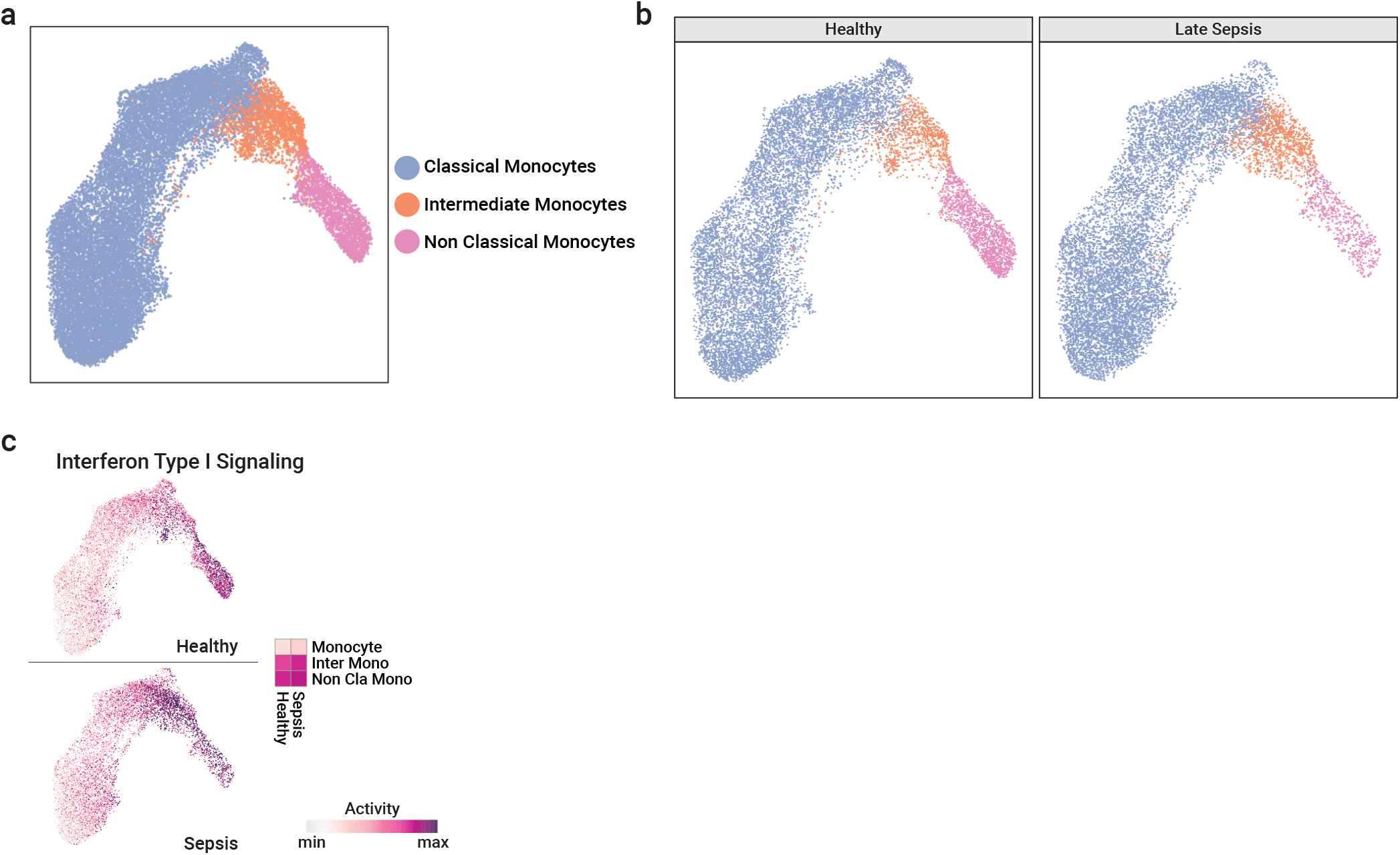
Loss of non-classical monocytes in patients with late sepsis. **a**, UMAP representation of blood monocytes obtained from late sepsis patients^21^, colored based on cell type. **b**, UMAP of late sepsis blood monocytes separated by health condition (healthy or late sepsis) and colored based on cell type. **c**, UMAP of IFN-I signaling pathway activity on late sepsis blood monocytes, separated by health condition.

**Extended Fig. 8:**
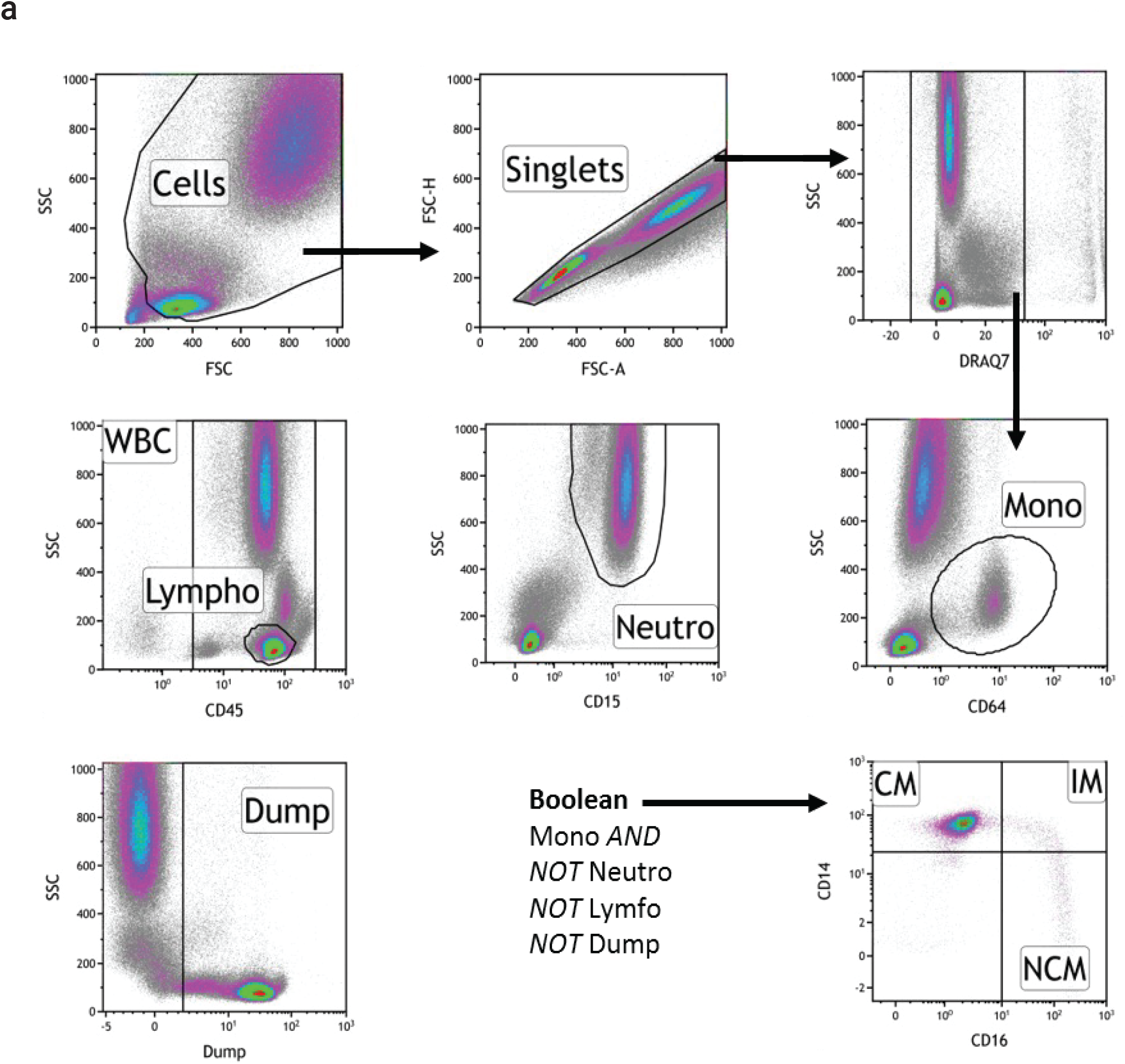
Gating strategy to analyze monocyte subtypes in blood. **a**, Gating strategy used to identify three monocyte subtypes in blood of LPS-SI volunteers by flow cytometry.

